# Indexing Cerebrovascular Health Using Near-infrared Spectroscopy

**DOI:** 10.1101/2021.05.09.21256921

**Authors:** Rashid Afkhami, Frederick Rohan Walker, Saadallah Ramadan, Rachel Wong, Sarah Johnson

## Abstract

Near-infrared spectroscopy (NiRS) is a relatively new technology of brain imaging with its potential in the assessment of cerebrovascular health only recently discovered. Encouraging early results suggest that NiRS can be used as an inexpensive and portable cerebrovascular health tracking device using a recently proposed pulse relaxation function (PReFx). In this paper, we propose a new NiRS timing index, TI_NiRS_, of cerebrovascular health. TI_NiRS_ is a novel use of the NiRS technology. TI_NiRS_ is motivated by the previously proved relationship of the timing of the reflected wave with vascular resistance and compliance in the context of pressure waveforms. We correlated both TI_NiRS_ and PReFx against age, a non-exercise cardiorespiratory fitness (CRF) index, and two existing indices of cerebrovascular health, namely Transcranial Doppler (TCD) augmentation index, AI_TCD_, and magnetic resonance imaging (MRI) blood flow pulsatility index, PI_MRI_. The TI_NiRS_ correlations with Age, CRF, PI_MRI_ and AI_TCD_ all are significant, i.e., *r* = 0.53 (*p* = 0.002), *r* = −0.44 (*p* = 0.011), *r* = 0.45 (*p* = 0.012) and *r* = 0.46 (*p* = 0.010), respectively. PReFx, however, did not have significant correlations with any of the vascular health factors. The proposed timing index is a reliable indicator of cerebrovascular aging factors in the NiRS waveform.

## Introduction

Vascular stiffening is an important health indicator and influences health in a manner that is independent of blood pressure [1]. The American Heart Association recommends that vascular stiffening be used as a predictor of cardiovascular disease risk [1]. Vascular stiffness is inversely proportional to compliance and reflects arterial resistance to stretching and recoiling in response to blood ejection from the heart. In a young or otherwise healthy vascular system, compliant arteries help smooth out the vascular pulsatility induced by blood pressure. As compliance decreases, blood pressure causes structural changes within the arterial walls that adversely affect their function [2, 3].

Another change that occurs with reduced vascular compliance is increased pulse wave velocity (PWV) [4]. PWV is the speed of the pressure wave traveling from the heart towards the periphery. Simultaneous measurement of arterial pulse in a proximal and a distal arterial sites e.g. common carotid and femoral gives the travel time of the pressure waveform. The distance between the measurement sites can be measured over the body surface. Hence, PWV is calculated as distance divided by travel time between two measurement sites [5]. PWV is commonly measured by applanation tonometry, however, other non-invasive technologies such as MRI and Doppler ultrasound can also be used [5]. A photoplethysmograph (PPG) which uses infrared light emitters and detectors to measure blood volume changes in periphery (e.g. fingertip, ankle, wrist or earlobe), is similarly used to estimate PWV by dividing subject height to the travel time [6, 7, 8].

When a pressure wave reaches a vascular bifurcation point, a portion of the wave is reflected back towards the heart. The reflected wave eventually augments an incident wave coming from the heart, thereby increasing the pulse pressure. Due to the increased PWV in a stiff arterial system, the reflected wave travels faster, thereby reaching the incident wave earlier in systole and increasing the local pulse pressure even further. Both forward and reflected waves leave a peak or an inflection point on the pressure waveform. The time difference between these peaks is believed to be an estimation of the travel time and used in calculation of PWV and PPG SI [9, 6, 10, 11].

Increased arterial stiffness can damage the arterial endothelium in small arteries and can, over time, produce a pathology known as small vessel disease (SVD) [2]. The brain, in particular, is highly vulnerable to SVD, as the blood flow to the sub-surface brain structures is compromised [12, 13, 14]. Based on a comprehensive review, cerebral SVD is believed to be one of the main causes of vascular cognitive impairments and accounts for a quarter of all acute ischaemic strokes [15].

Cerebrovascular compliance is commonly assessed non-invasively using indices derived from blood pressure and flow readings. Transcranial Doppler (TCD) ultrasound and magnetic resonance imaging (MRI) are the modalities most widely used to acquire cerebrovascular compliance and cerebrovascular health indices. TCD measures blood flow velocity; i.e., the speed at which the blood cells move inside a vessel. While it is possible to estimate compliance using TCD blood flow velocity [16], the method relies on estimates of vascular cross-sectional area, arterial inflow and venous outflow, which are not readily measurable. The TCD indices such as the pulsatility index, PI_TCD_, and augmentation index, AI_TCD_, are the commonly reported TCD cerebrovascular health indicators [17, 18, 19]. Of these two, PI_TCD_ is the most commonly used TCD index due to its ease of calculation. However, AI_TCD_ has been shown to have a stronger correlation with cerebrovascular health indicators such as age and cardiorespiratory fitness (CRF) [20]. The MRI pulsatility index (PI_MRI_) is the commonly adopted MRI-based cerebrovascular health measure [14]. While MRI has a high spatial resolution for localized measurements, it is less commonly used as it is both costly and time consuming, making it unsuitable for simple and routine monitoring of vascular health in the general population.

Near-infrared spectroscopy (NiRS) is a relatively new technology that measures regional blood volume changes based on absorption of near-infrared light, mainly by oxygenated blood inside cerebral arteries [21, 22, 23]. NiRS offers high temporal resolution and is sufficiently sensitive to detect changes in blood volume during the cardiac cycle. Therefore, NiRS allows local studies of cerebrovascular behaviour and possible assessment of cerebrovascular health [21]. The recently proposed pulse relaxation function (PReFx) is the first NiRS based approach to the indexing of cerebrovascular compliance [21]. PReFx measures the deformation of the blood volume waveform during the systolic relaxation phase due to the presence of reflected waves. It has been demonstrated to correlate with CRF and inversely correlate with age [21, 24, 25]. PReFx is an area measurement in the averaged NiRS signal. The PReFx algorithm assumes that the first positive peak after the diastolic minimum is the systolic peak, which may not be the case when the systolic maximum is an inflection point rather than a peak. Therefore, we are motivated to use an index that can be easily applied to NiRS signals. In our previous works we have proposed a new timing index (TI) which which has been shown to significantly correlate with vascular health factors such as age and CRF in a mathematical model of the pressure wave [11] and in an experimental study using TCD waveforms [20]. TI is defined as the inverse of the reflection time, i.e. the time difference between the systolic and reflected peaks, and using a tube-based model of the arterial tree, the timing index has been proven to be directly controlled by vascular aging factors including compliance, resistance and PWV [11]. In fact, it has been shown that the reflection time is more than the time it takes for the blood waveform to travel the distance between the measurement site and the reflection site and the waveform faces an additional delay before turning back from the reflection site which is a function of resistive and capacitive properties of the vascular system beyond the reflection site [11]. Therefore, the reflection time is a sum of two time delays, one is a line delay controlled by PWV and physical distance between the measurement and reflection sites and the other is a load delay forced by the vascular properties beyond beyond the reflection site. All these parameters are known to change with ageing and vascular health and thus making TI a powerful vascular stiffness index [11]. As such, we propose a new NiRS timing index (TI_NiRS_) to directly measure the timing of reflected waves. Here, the NiRS timing index is defined as the inverse of the time between the systolic and reflected waveform peaks in the NiRS signal. Then, we will examine the correlation of the proposed index with other health indices including age, CRF and the most currently used vascular health measures derived from other imaging modalities, namely TCD augmentation index and MRI pulsatility index.

## Methods

### Protocol and Data Collection

Thirty eight adults (23 female, 15 male, age range = 24-67 years, mean age = 41.7 years) were recruited from the Newcastle, Australia. They provided informed consent prior to assessment. The study protocol was approved by the University of Newcastle Human Research Ethics Committee and is registered in the Australian and New Zealand Clinical Trials Registry (ACTRN12619000144112). All experiments were performed in accordance with relevant guidelines and regulations. TCD recordings from these participants were previously reported in [20]. Each participant attended the Hunter Medical Research Institute and the University of Newcastle Callaghan campus over two consecutive days and participated in three scanning sessions. Participants were asked to refrain from consuming caffeine before their scans. Height, weight, age, sex and resting heart rate were recorded for each participant and a physical activity questionnaire was completed.

TCD ultrasound (DopplerBox X; Compumedics DWL, Singen, Germany) was used to record resting-state cerebral blood flow velocity from the right and left middle cerebral arteries. Participants were also scanned on a 3T MRI scanner (Magnetom Prisma, Siemens Healthineers, Erlangen, Germany), equipped with 64-channel receive only head coil, while a standard built-in dual channel body coil was used for RF transmission. Blood flow was quantified using a phase contrast flow quantification sequence (TR = 26.5ms, TE = 6.9ms, slice thickness = 5mm, matrix 256 *×* 256) of a single excitation with a velocity encoding value of 120cm/s on middle cerebral arteries. Finally, participants were instructed to relax and sit still for 300 seconds of resting-state NiRS recording. A frequency-domain NiRS (ISS Imagent, Champaign, Illinois, USA) was used at a 110MHz modulation frequency and sampling rate of 39.0625Hz. The setup montage was designed to cover the majority of the frontal lobe and consisted of four detectors, each crossed with 16 time-multiplexed sources (eight of which operated at 690nm and the other eight at 830nm), making a total of 64 channels. Participants were asked to wear an Equivital EQ02 LifeMonitor sensor belt during this session, which recorded ECG and NiRS signals simultaneously.

During the TCD imaging session, resting-state heart rate was measured using an heart rate monitoring device (Omron HEM-7320). Participants assumed a sitting position for approximately five minutes during headpiece setup before recording commenced. During TCD recording, the heart rate was measured once per minute three times. The mean of these three heart rate measurements was used as the resting-state heart rate.

### Calculating the Indices

#### MRI Pulsatility Index

MRI phase contrast images were processed using the scanner software (Siemens Syngo) by manually placing a region of interest around the middle cerebral artery and quantifying the flow. The MRI pulsatility index, PI_MRI_, was then obtained by subtracting the minimum flow from the maximum flow and dividing it by the mean flow over the cardiac cycle [14]; i.e.,

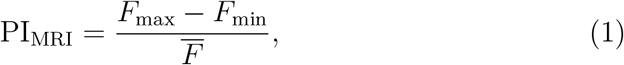

where *F* indicates the flow and the bar sign indicates the mean value. Three subjects did not have an MRI scan and among the remaining 35 subjects, PI_MRI_ was calculated either for both middle cerebral arteries (*n* = 21) or for a single middle cerebral artery (MCA) (*n* = 14) based on data quality. The indices from right and left MCAs were then averaged for each subject.

#### TCD Augmentation Index

Calculation of the augmentation index, AI_TCD_, requires identification of three characteristic points on an averaged Doppler waveform. Peak systole (*V*_sys_), peak diastole (*V*_dia_) and the peak of the reflected wave (*V*_refl_) are the parameters required to calculate the augmentation index [18]:

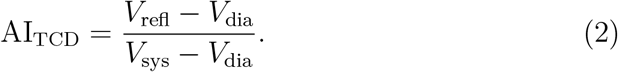

Thus, the timing of the diastolic peaks was first determined automatically in MATLAB using a peak detector function. Then, each signal was averaged with respect to its diastolic peaks, resulting in an averaged signal covering the duration of a single cardiac cycle. The systolic and reflected peaks were set manually on the averaged signal. The TCD data for two subjects were excluded from the analysis due to noise. The AI_TCD_ index was calculated using data from both MCAs (and averaged) for 34 subjects and from a single MCA for two subjects, depending on data quality.

#### Evaluating Cardiorespiratory Fitness

The CRF was determined using the non-exercise test model proposed in [26]. It involves a self-reported physical activity score, age, sex, body mass index and resting heart rate. See the *supplementary materials* for the physical activity questionnaire and cardiorespiratory fitness formula.

#### NiRS Indices

Optical light intensity NiRS signals were used to derive two different indices: the pulse relaxation function (PReFx) and the proposed timing index, TI_NiRS_. The pre-processing of the recordings was done similarly for both methods and was adopted from [21] using the “*p pod*” package [27] which is a MATLAB-based software module developed by the same group. Specifically, channels with source detector distances of 20-60 mm were selected and went through a quality-control process passing 1283 channels out of the original 38 *×* 64 = 2432 channels. These channels were then normalized by dividing by their mean values and then bandpass-filtered to 0.5-5.0 Hz to capture the dynamics of the pulse signal and remove any low-frequency elements and moving artefacts. The resulting signal was averaged with respect to the ECG R peaks, and then rotated around the x-axis (see Fig. 1). The result is called the arterial pulsation signal, *s*. The rotation was performed so that the systolic peak would appear as a positive peak and the shape of the signal would look similar to that of a blood pressure, flow or volume signal. This facilitates understanding of the waveform. The arterial pulsation signals were visually inspected and channels that did not have a pulse shape were removed (i.e., low blood volume at the beginning and end representing diastole, and an increase in between representing systole). Some 964 channels passed the pre-processing stage for further analysis. Channels with source detector distances between 25 and 50 mm proved to be most likely to generate usable waveforms.

**Figure 1:**
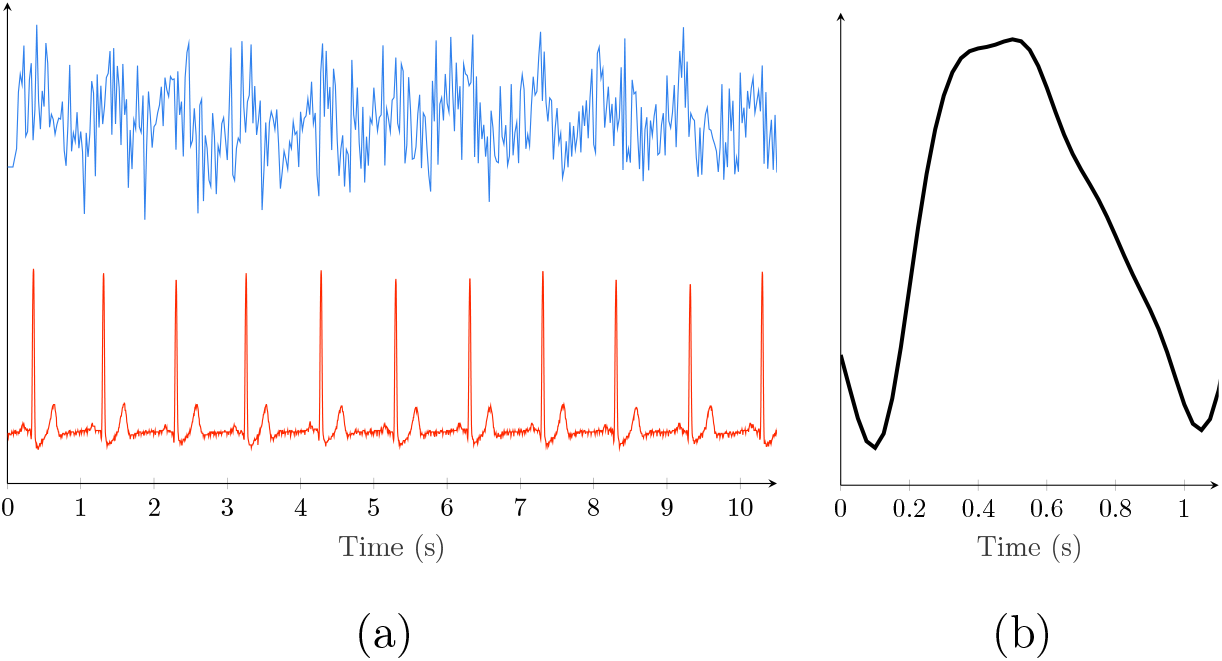
(a) Ten-second-long raw NiRS signal (blue) and simultaneously recorded ECG lead I (red). (b) Averaged and flipped NiRS *s* signal.

#### PReFx

The pulse relaxation function is calculated based on the methods explained in [21]. In brief, *s*_D1_ and *s*_D2_ peaks occur as minima in the *s* signal in the first and the second cardiac cycles and *s*_S_ occurs as the first local maximum after *s*_D1_ (see Fig. 2). {*s*_D1_, *s*_D2_} and *s*_S_ are intended to represent the diastolic and systolic peaks, respectively. However, *s*_S_ is not necessarily a systolic peak, see Fig.2b where *s*_S_ is the reflected peak and the systolic peak indicated by an arrow. Then, the area enclosed by the *s* signal between *s*_S_ and *s*_D2_ is calculated as *A* and inserted into

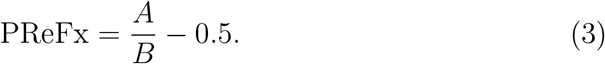

**Figure 2:**
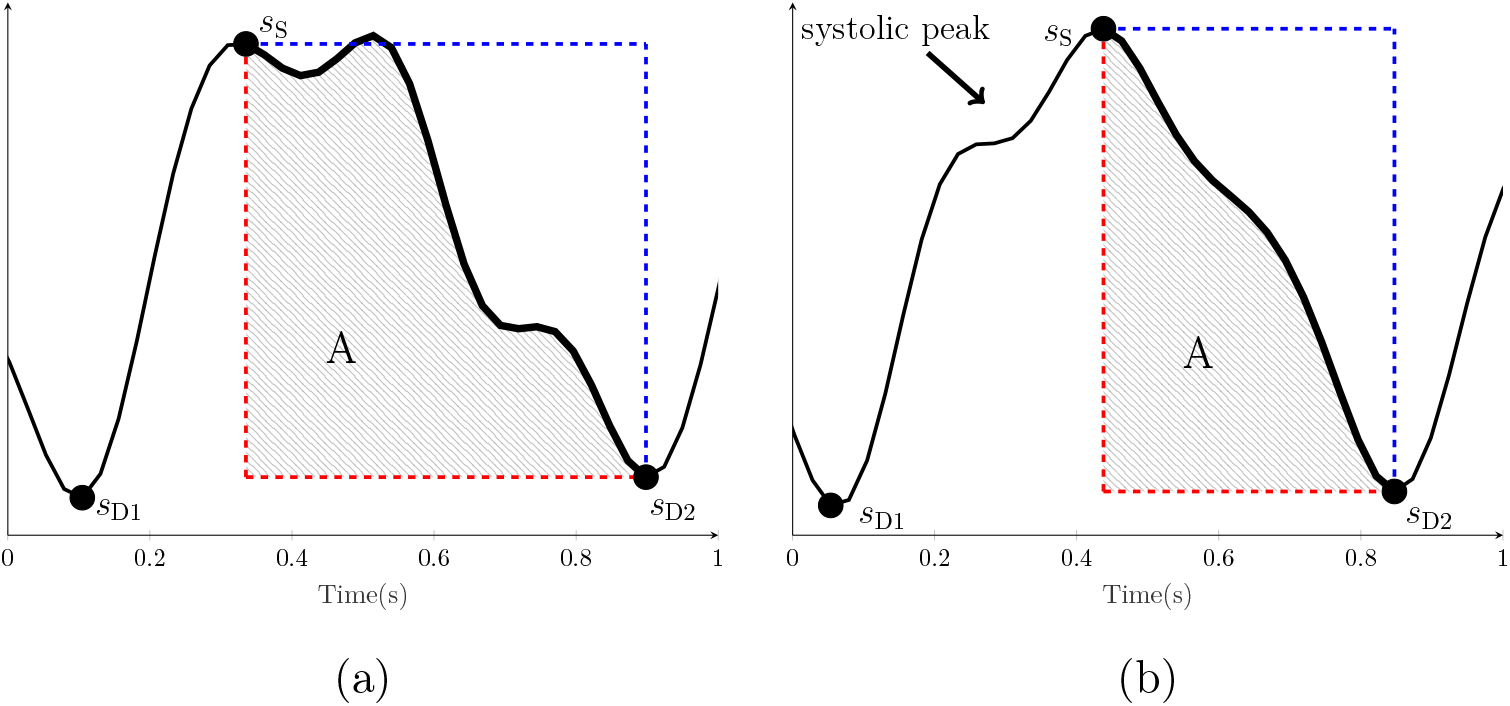
Examples of calculating PReFx for two different *s* signals. The signal *s* is shown in black, the *s*_S_, *s*_D1_ and *s*_D2_ peaks are marked with black circles, and the blue and red rectangles are formed with *s*_S_ and *s*_D2_ at their diagonals. The shaded area, *A*, is the area enclosed by *s* between *s*_S_ and *s*_D2_ and the red sides of the rectangle. The calculated PReFx values for (a) and are 0.136 and 0.081, respectively.

Where *B* is the area of a rectangle formed by *s*_S_ and *s*_D2_ at its diagonal ends (i.e. the rectangle formed by blue and red lines in Fig. 2). Note that if the drop in volume between peaks was linear, PReFx would equal zero. After communicating with the authors of [21], channels with PReFx value outside the interval of [−0.1, 0.4] were removed and other channels were manually excluded from the calculations where reflected peaks were selected by the algorithm (see Fig. 2b for an example). A total of 245 additional channels were removed in this process. Finally, for each subject with more than ten PReFx values, an averaged PReFx value was calculated as the index of vascular compliance. We were able to calculate PReFx for 29 of the 38 subjects.

#### TI_NiRS_

We define the timing index as the inverse of the time between the forward and reflected waveform peaks. In order to find the timing of these peaks, named *t*_sys_ and *t*_refl_ for peak systole and peak reflection, respectively, for a given signal, *s*, a completely automated algorithm is proposed (Sample data and codes available at https://github.com/Rashid-Afkhami). The algorithm uses the second derivative to locate key points in the signal, this is based on similar approaches that have been proposed for locating inflection point in the pressure waveform [28]. Firstly, the second derivative, *s*_2_, of *s* is calculated. Next, the following time points are defined on the *s* and the *s*_2_ signals: {*t*_p1_, *t*_p2_, …,} the time of the peaks on the *s* signal and {*t*_zc1_, *t*_zc2_, …} the time of zero-crossings on the *s*_2_ signal. If the number of *s*_2_ zero-crossings is less than 4, the channel is excluded. Otherwise,

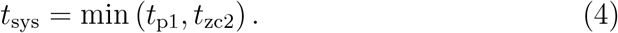

Then, to define *t*_refl_ the algorithm checks only the time period between *t*_zc3_ and *t*_zc4_. If *s* has a positive peak, *t*_refl_ is set as the time point corresponding to this peak, *t*_pi_, otherwise, it is set as the time when *s*_2_ reaches its minimum, *t*_min_ (all in the specified time period between *t*_zc3_ and *t*_zc4_) i.e.,

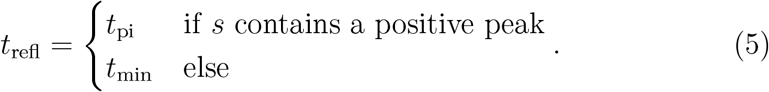

After all channels are processed for a subject, channels with *t*_sys_ and *t*_refl_ outside their mean *±*1.5 std (i.e., standard deviation) or outside a predefined threshold were also excluded from further calculations. The predefined threshold for *t*_sys_ was any value <125 ms and for *t*_refl_ it was any value >500 ms. Then, if ten or more channels remained, the *t*_sys_ and *t*_refl_ values for each channel were used to calculate the timing index as:

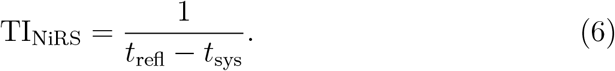

The timing index values of different channels were then averaged for each participant. Of the original 38 subjects, we were able to calculate a TI for 32 of them. Examples of the proposed algorithm are shown in Fig. 3 when the algorithm is applied to in vivo and simulated signals. Three main types of the arterial pulsation signals, *s*, are shown in this figure where the detected *t*_sys_ and *t*_refl_ correspond to either a peak point or an inflection point on the recorded signal. The simulated signals are formed by adding three waveforms, an incident forward-traveling waveform, a reflected waveform and a re-reflected waveform (which is also traveling towards the periphery). The purpose of the simulated waveforms is to better visualize where the underlying waveforms reach their peak and how the algorithm performs in terms of detecting the peak times. For each simulated case the underlying waveforms are arranged in a form that the resulted observed signal (black line) resembles an in vivo case. Using a NiRS device only the composite signal *s* is detected and the wave components shown in Fig. 3b, d and f are for illustrative purposes only.

**Figure 3:**
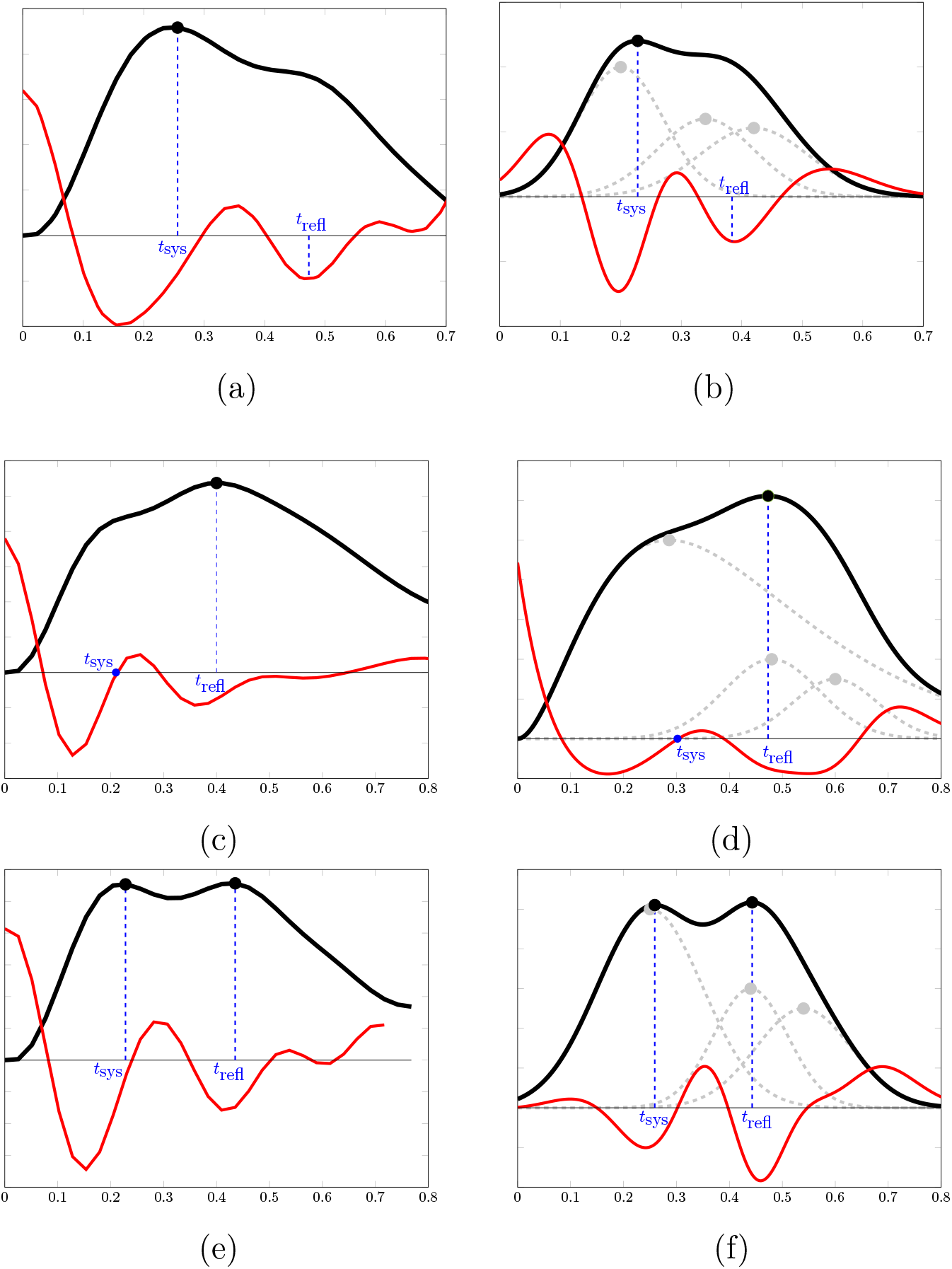
Three main types of in vivo arterial pulsation signals are shown on the left column (a), (c) and (e) and an equivalent simulated signal is shown on the right column (b), (d) and (f), respectively. Black lines are the observed recorded blood volume signal, *s* and the red lines are the second derivatives, *s*_2_. The gray lines represent the underlying forward, reflected and re-reflected waveforms. The x-axis represents time in seconds.

## Statistical Analysis

Statistical analysis was carried out in MATLAB using the inbuilt *corr* function. Lilliefors test of normality with a 5% significance level was used to assess the data distribution. Age and PI_MRI_ were found to be non-normally distributed (*p* = 0.0072 and 0.0020, respectively). All other distributions were normal. Pearson or Spearman correlation coefficients were calculated for normally-distributed and non-normally-distributed data, respectively. False discovery rate correction was used to adjust significance levels with an initial *α* = 0.05.

## Results

The correlations of the proposed NiRS timing index and existing NiRS PReFx index with the indices derived from other imaging modalities, age and CRF are reported in Table 1. Corresponding scatter-plots are shown in Fig. 4. Least-squares linear models have not been provided for Fig 4.G and Fig 4.H due to small slopes. Significant correlations after correcting for multiple comparisons are marked with an asterisk. Note that PReFx is an index of vascular compliance whereas TI_NiRS_ is an index of vascular stiffness that is inversely proportional to compliance. Therefore, the two indices show opposite signs when correlated with the same factors. The proposed systolic peak and reflected wave peak detection algorithm for TI_NiRS_ detected 228 channels (out of 964 input channels) with a systolic peak corresponding to an inflection point (mean participant age for these channels = 42.5 years). Both TI_NiRS_ (the peak detection algorithm) and PReFx indices are affected by the sampling frequency of 39.0625Hz (i.e. *≈* 25ms sampling interval) used in this study; a faster sampling rate would allow a higher resolution and more accurate results.

**Table 1:** Correlation matrix with correlation coefficients (and *p*-values). Asterisks indicate statistically significant correlations after correction for multiple comparisons using a false discovery rate

|  | CRF | Age | AI<sub>TCD</sub> | PI<sub>MRI</sub> |
| --- | --- | --- | --- | --- |
| TI<sub>NIRS</sub> | -0.44* (0.011) | 0.53* (0.002) | 0.46* (0.010) | 0.45* (0.012) |
| PReFx | 0.24 (0.203) | -0.40 (0.030) | -0.26 (0.173) | -0.01 (0.978) |

**Figure 4:**
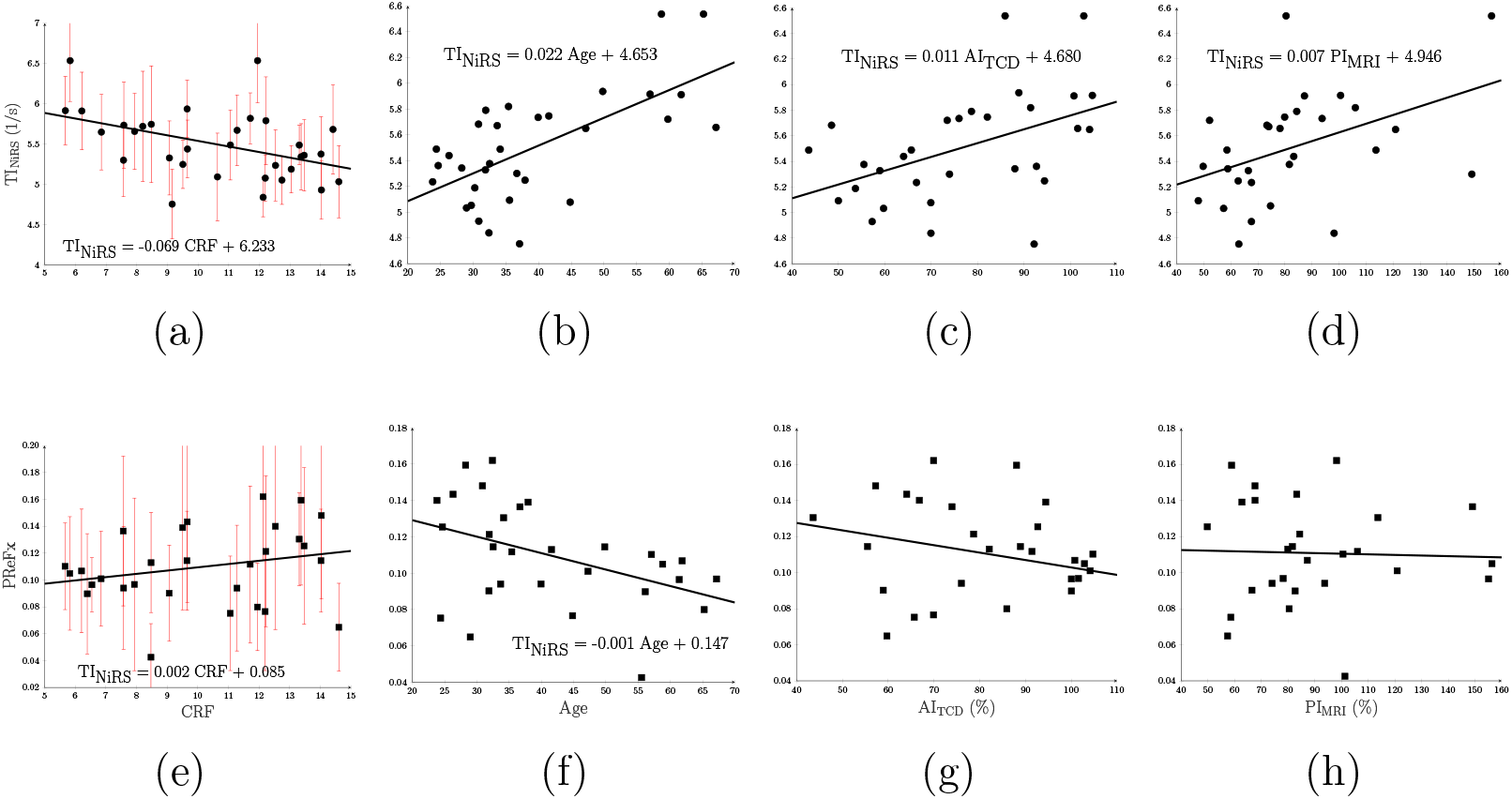
Scatter plots of the correlations with least-squares fit line. (a), (b), and (d) represent TI_NiRS_ scatter plots with CRF, age, AI_TCD_ and PI_MRI_, respectively. (e), (f), (g) and (h) represent PReFx scatter plots with CRF, age, AI_TCD_ and PI_MRI_, respectively. The red bars in (a) and (e) show TI_NiRS_ and PReFx standard deviations, respectively.

## Discussion

The NiRS indices are designed to measure vascular stiffness, which is well correlated with age [1] and cardiorespiratory fitness [29]. Thus, a common approach with any vascular index is to compare its relationship with age and CRF [30, 21] and here we have done the same. The PReFx correlation coefficient reported here for age is consistent with existing studies; however, the correlation with CRF is lower than previously reported. PReFx correlations with age and CRF in the adult population have so far been studied by the same research group using two different datasets [21, 24]. Reported correlations with age are *r* = −0.39 (mean age = 69.87 years) [21] and *r* = −0.43 (mean age = 47.8 years) [24]; a correlation of *r* = −0.60, mean age = 58.8 years was reported in [25] for a combination of the two datasets. Our reported correlation, *r* = *—*0.40 (*p* = 0.030), although consistent with the previous values, is not significant. Reported correlations with CRF are *r* = 0.416 [21] and *r* = 0.32 [24], however, our correlation with CRF of *r* = 0.24 (*p* = 0.203) was not significant. Given that the mean age for our sample is 41.7 years, we suspect that the PReFx measure has a lower correlation in younger subjects than older ones. Based on the performance of the PReFx algorithm on the current data, we have identified two potential sources that can distort the PReFx calculation and potentially cause lower correlations. Firstly, there are cases where the PReFx algorithm detects reflected peaks instead of systolic peaks as the *s*_S_ points. The PReFx algorithm assumes the first local maximum after the minimum peak, i.e. *s*_S_, to be the systolic peak, which is not accurate when the true systolic peak is in the form of an inflection point (e.g., see Fig. 2b and 3b). Based on our analysis these cases account for almost one-fourth of all channels (228 of the 964 studied channels) and are more likely to occur in older participants in which the reflected wave moves faster and reaches the incident wave sooner than in younger participants. The second problem occurs in channels with a notch in the signal after the systolic and reflected peaks, which is likely caused by closure of the aortic valve (similar to the dicrotic notch seen in the pressure waveform [31]). This notch can appear as a large deformation in the signal (see Fig. 2a for an example), which will change PReFx values independently of vascular health factors. Among the aforementioned two potential causes of low correlation for PReFx, the first was eliminated in the results reported in Table 1 by discarding channels where *s*_S_ was assigned to the reflected peak. However, if all the channels are included in the calculations, it will remove the significant correlation of PReFx with age; i.e., *r* = −0.19.

While no previous NiRS studies have reported a timing index, the correlations of TI_NiRS_ with age and CRF match those of the TCD reported by our group [11]. That is, correlations of *r* = 0.70 and *r* = 0.50 between TCD TI and age in two different datasets and a correlation of *r* = −0.79 between TCD TI and CRF. The proposed NiRS timing index, TI_NiRS_, also shows significant correlations with age (*r* = 0.53, *p* = 0.002) and CRF (*r* = −0.44, *p* = 0.011). TI_NiRS_ correlated more strongly with both age and CRF than PReFX. This suggests that TI may be a more sensitive index than PReFX to age and lifestyle related cerebrovascular changes in middle-aged participants.

Based on a previously proven association between PI_MRI_ and SVD [14], we expect NiRS indices to correlate with the MRI pulsatility index which is an indicator of vascular health. PReFx showed no significant correlation with PI_MRI_ (*r* = −0.01, *p* = 0.978), whereas TI_NiRS_ correlated significantly with PI_MRI_ (*r* = 0.45, *p* = 0.012). PI_MRI_ indicates vascular health by measuring the pulsatility of the blood flow. High pulsatility means that fast traveling reflected waves join incident waves and contribute to increased maximum flow peaks. Similarly, TI_NiRS_ reaches greater values when the reflected wave peak is closer to the systolic peak and pulsatility is high. PReFx also changes based on the location of the reflected wave; high PReFx values correspond to channels with high deformation from a perfect sinusoidal waveform, i.e., the existence of a reflected peak. However, the presence of a dicrotic notch may also affect the PReFx index by distorting the wave shape while having no effect on PI or TI. Thus, in channels with a prominent dicrotic notch the PReFx may be high while pulsatility is low, which may explain the lower correlation for PReFx.

TI_NiRS_ showed a significant correlation with the TCD augmentation index (*r* = 0.46, *p* = 0.010), while the correlation of PReFx with AI_TCD_ was not significant (*r* = −0.26, *p* = 0.173). The augmentation index is a widely accepted index used in both blood pressure and flow velocity (TCD) signals, and is a strong predictor of vascular aging and stiffening [32, 18, 19, 33]. The AI captures the augmentation of the incident wave by the reflected wave, with smaller values reflecting no augmentation when the arteries are healthy and the propagation speed is low. TI indexes this delay between the incident and reflected waves directly, which is why AI_TCD_ and TI_NiRS_ are significantly correlated. The PReFx also correlates with AI as larger PReFx values can correspond to late reflected waves leaving a peak on the systolic-to-diastolic segment of the arterial pulsation signal and thus increasing the area under the curve. Nevertheless, the correlation may be weakened when the presence of dicrotic notch increases PReFx regardless of the reflected wave.

Overall, NiRS blood volume waveforms are believed to closely resemble the changes in blood pressure [21] which is also supported by the use of infrared light in PPG and its morphological similarity to the arterial blood pressure waveform [22, 34]. Thus, we believe that the proposed NiRS timing index is affected similarly by vascular health parameters in the same way as the timing index derived from the blood pressure waveforms [11]. As blood pressure can not be readily measured in cerebral vessels, a NiRS-based timing index can provide an alternative approach.

## Conclusion

In this paper, we presented a novel NiRS based cerebrovascular stiffness index, TI_NiRS_. The TI_NiRS_ correlates significantly with age, CRF and other cerebrovascular health indices derived from TCD and MRI data, indicating it has high performance in tracking changes in the cerebrovascular system. NiRS offers a potentially valuable means of indexing vascular health and has superior cost, portability and widespread implementation potential compared with existing techniques.

## Data Availability

The datasets generated during and/or analysed during the current study are available from the corresponding author on reasonable request.

