## Supplementary Materials for "Indexing Cerebrovascular Health Using Near-infrared Spectroscopy"

**Estimation of CRF**

Methods based on [Jurca et al., 2005]

**Step 1:** Acquiring physical activity score

| **Physical Activity Description** | **Score** |
|---|---|
| Inactive or little activity other than usual daily activities | 0.00 |
| Regularly ($\geq$5 d/wk) participate in physical activities requiring low levels of exertion that result in slight increase in breathing and heart rate for at least **10 minutes** at a time | 0.32 |
| Participate in aerobic exercises such as brisk walking, jogging or running, cycling, swimming, or vigorous sports at a comfortable pace or other activities requiring similar levels of exertion for **20 to 60 minutes** per week | 1.06 |
| Participate in aerobic exercises such as brisk walking, jogging or running at a comfortable pace, or other activities requiring similar levels of exertion for **1 to 3 hours** per week | 1.76 |
| Participate in aerobic exercises such as brisk walking, jogging or running at a comfortable pace, or other activities requiring similar levels of exertion for **over 3 hours** per week | 3.03 |

**Step 2:** Estimating CRF

$$\begin{aligned} \text{CRF} = &+ 2.77 \times (\text{0 for women, 1 for men}) \\ &- 0.10 \times (\text{Age in years}) \\ &- 0.17 \times \left(\text{Body mass index in kg/m}^2\right) \\ &- 0.03 \times (\text{Resting heart rate in beats per minute}) \\ &+ 1.00 \times (\text{Physical activity score}) \\ &+ 18.07 \end{aligned}$$

[Jurca et al., 2005] Jurca, R. et al. (2005). Assessing cardiorespiratory fitness without performing exercise testing. *American Journal of Preventive Medicine*, 29(3):185–193.
